# Enrichment of the local synaptic translatome for genetic risk associated with schizophrenia and autism spectrum disorder

**DOI:** 10.1101/2023.10.19.23297263

**Authors:** Nicholas E Clifton, Julie Qiaojin Lin, Christine E Holt, Michael C O’Donovan, Jonathan Mill

**Affiliations:** Department of Clinical & Biomedical Sciences, Faculty of Health and Life Sciences, University of Exeter, Exeter, UK; UK Dementia Research Institute, Department of Clinical Neurosciences, University of Cambridge, Cambridge, UK; UK Dementia Research Institute, King’s College London, London, UK; Department of Physiology Development and Neuroscience, University of Cambridge, Cambridge, UK; Division of Psychological Medicine and Clinical Neurosciences, Cardiff University, Cardiff, United Kingdom

**Keywords:** Local translation, synapse, schizophrenia, autism spectrum disorder, genome-wide association study, transcriptome

## Abstract

**Background:** Genes encoding synaptic proteins or mRNA targets of the RNA binding protein, Fragile X mental retardation protein (FMRP), have been linked to schizophrenia and autism spectrum disorder (ASD) through the enrichment of genetic variants conferring risk to these disorders. FMRP binds many transcripts with synaptic functions and is thought to be a key regulator of their local translation, a process which enables rapid and compartmentalized protein synthesis required for development and plasticity.

**Methods:** Here, we used summary statistics from large-scale genome-wide association studies to test the hypothesis that the subset of synaptic genes encoding localized transcripts is more strongly associated with schizophrenia and ASD than non-localized transcripts. We also postulated that this subset of synaptic genes is responsible for associations attributed to FMRP targets.

**Results:** We show that schizophrenia associations were enriched in genes encoding localized synaptic transcripts compared to the remaining synaptic genes, or to the remaining localized transcripts; this also applied to ASD associations, although only for transcripts observed after stimulation by fear conditioning. The genetic associations with either disorder captured by these gene sets were independent of those derived from FMRP targets. Furthermore, we found that schizophrenia association was related to FMRP interactions with mRNAs in somata, but not in dendrites, whilst ASD association was related to FMRP binding in either compartment.

**Conclusions:** Our data suggest that synaptic transcripts capable of rapid and compartmentalized local translation are particularly relevant to the pathogenesis of schizophrenia and ASD, but do not characterize the associations attributed to current sets of FMRP targets.

## Introduction

Common neuropsychiatric and neurodevelopmental disorders are leading causes of disability among young adults and many cases remain poorly treated by current medications (1). Advances in psychiatric genetics (2–6) have highlighted regions of the genome, and specific genes, associated with risk for neuropsychiatric disorders, yet our understanding of the cellular mechanisms through which they confer risk has been insufficient to effectively target new therapies. To reach a point where we can improve treatments, there is a need to refine the biological context in which genetic risk converges on common pathways, taking into account the dynamic and compartmentalised nature of neuronal processes.

Genomic and functional evidence implicates the molecular machinery responsible for synaptic function and plasticity in the pathophysiology of schizophrenia and autism spectrum disorders (ASD) (7–11). Synaptic plasticity is a time-sensitive process, occurring in response to localized extrinsic stimuli. An important requirement of synaptic plasticity is the ability of cells to regulate the maturation and strength of individual synapses quickly and independently of other synapses from the same cell, a process that is facilitated by local synthesis of new proteins in specific neuronal compartments undergoing plasticity (12). The RNA binding protein (RBP), Fragile-X mental retardation protein (FMRP), is considered to be important for this process, as it plays a key role in regulating both the transport and activity-dependent local translation of many transcripts required for synaptic development and plasticity (13, 14). Loss of FMRP function is a monogenic cause of developmental disorders, including ASD, and the transcripts bound by FMRP are enriched for genetic variation associated with both schizophrenia (15, 16) and ASD (14, 17–20). Furthermore, cytoplasmic FMRP-interacting protein 1 (CYFIP1), which forms a complex with FMRP and the translation initiation machinery to repress translation, has also been linked to both schizophrenia and ASD through copy number variant (CNV) deletions at 15q11.2 (21, 22). Collectively, these findings highlight the importance of studying local synaptic gene translation, and specifically, of investigating the relative contributions to risk of schizophrenia and ASD of genes translated locally, compared to those translated prior to transport to the synapse.

Whilst genetic variants associated with schizophrenia and ASD show a degree of pleiotropy (3, 23–25), the age of onset and clinical presentation of each disorder differs. One way these differences may originate is through variation in the effects of risk alleles on transcripts that influence synaptic plasticity during brain development, learning and memory, many of which may be locally translated. To evaluate the contribution of locally translated transcripts in driving the association of genes encoding synaptic proteins with schizophrenia and ASD, we used published, *in vivo* subcellular transcriptome and translatome datasets to classify synaptic genes by subcellular localization, and test their relationship with genetic variation. Second, with the aim of better characterising the schizophrenia association attributed to FMRP-regulated transcripts specifically, we examined the overlap between association signals derived from the local synaptic translatome and that from FMRP targets.

## Methods and Materials

### Gene sets

#### Synaptic gene ontology

Synaptic gene definitions were taken from manually curated functional annotations provided by the SynGO consortium (26). 1089 genes annotated to “synapse” were filtered to exclude those with a non-traceable author statement, leaving 1016 genes used for analysis and referred to herein as SynGO:*synapse*. These were subdivided into postsynaptic and presynaptic genes using SynGO annotations “postsynapse” and “presynapse”, consisting of 624 genes and 536 genes, respectively, with 236 genes being annotated to both compartments. A comparison set of synaptic gene annotations was obtained from the Gene Ontology (GO) database (27) (GO:0045202). After removing gene annotations with evidence codes NAS (non-traceable author statement), IEA (inferred from electronic annotation), or RCA (inferred from reviewed computational analysis), 611 genes remained for analysis, referred to as GO:*synapse*. Between SynGO:*synapse* and GO:*synapse* gene sets, 384 genes overlap.

#### Localized transcripts

Transcripts localized to synaptoneurosomes (fractionated synaptic terminals containing pre- and postsynaptic machinery) from mouse cortex at postnatal day 21 were obtained from Ouwenga et al (28). The local transcriptome was defined as transcripts enriched in synaptoneurosomes compared to the whole cell homogenate, with a false discovery rate (FDR) < 0.01. Mouse Ensembl IDs for 3408 genes encoding these transcripts were converted to human Ensembl IDs using Bioconductor biomaRt (29), for downstream analysis. Following removal of genes with zero or multiple human homologs, 3199 genes remained.

We obtained a set of ribosome-bound transcripts enriched in dendrites from adult (6-10 weeks) mouse hippocampal CA1 pyramidal neurons compared to ribosome-bound transcripts in cell bodies of the same set of neurons (30). The translatome was captured using compartment-specific translating ribosome affinity purification (TRAP) in conditionally tagged mice with RiboTag expression driven by *Camk2a*-Cre, directing the ribosome tag to CA1 pyramidal neurons. 1211 mouse gene symbols were converted to 1147 human Ensembl IDs, as above.

Thirdly, ribosome-bound transcripts from dendrites of adult (2-3 months) mouse hippocampal CA1 pyramidal neurons following exposure to a contextual fear conditioning trial were obtained from Ainsley et al (31). The dendritic translatome was extracted using TRAP with epitope-tagged ribosomal proteins driven by a *Camk2a* promotor through a tetracycline-controlled transactivator system. Mouse gene symbols, representing 1890 unique mRNAs enriched in dendritic ribosomes following fear conditioning compared to samples from home caged mice, were converted to 1577 human Ensembl gene IDs, after filtering, for analysis. For comparison, we obtained a second set of 2903 mRNAs bound to ribosomes in the soma after fear conditioning, from the same study. These were converted to 2442 human Ensembl gene IDs.

By intersecting synaptic functional annotations (SynGO:*synapse* or GO:*synapse*) with synaptoneurosome or dendrite transcriptomes or translatomes (Figure 1A; Supplementary Figure 1A), we created, for each intersection, three gene sets for comparison in genetic association analyses: synaptic genes translated locally; synaptic genes not translated locally; and non-synaptic genes translated locally.

**Figure 1.**
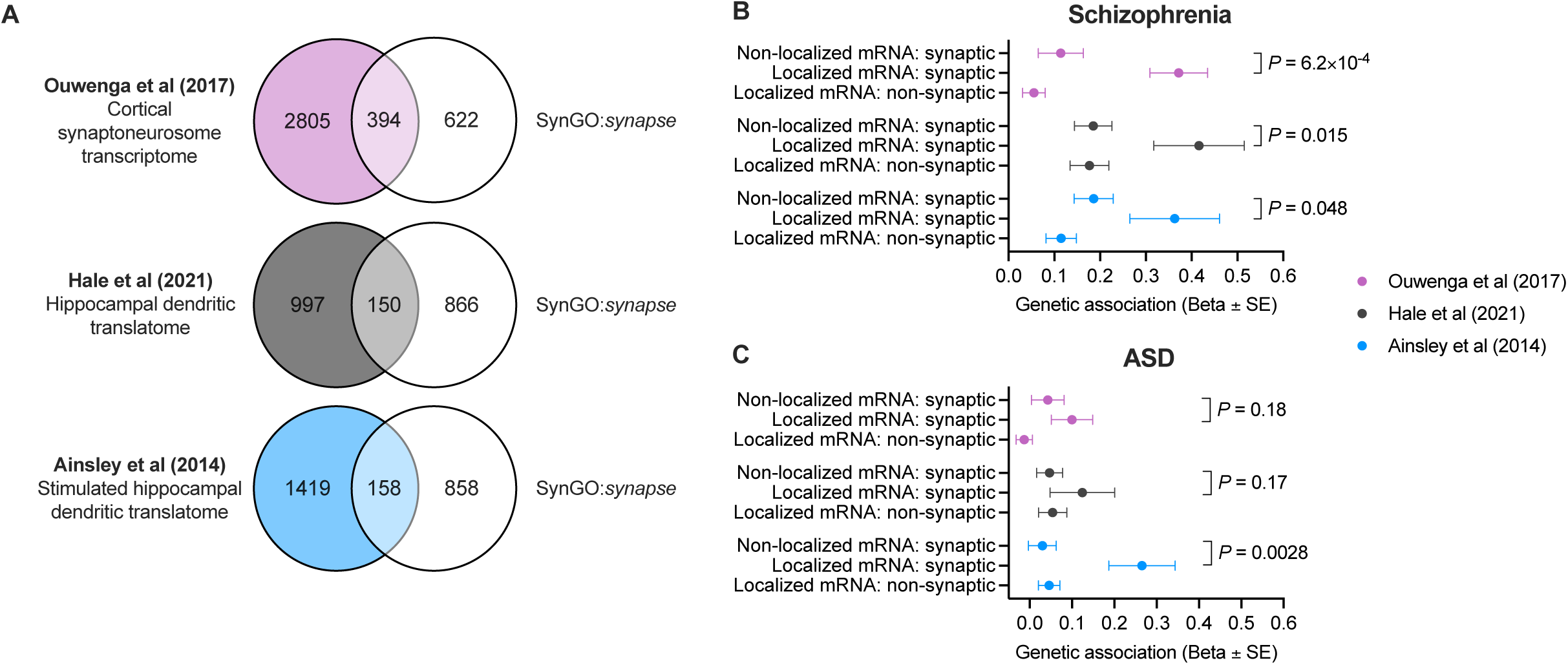
**A**. Intersection of localized mRNA transcripts with SynGO:*synapse* annotations. **B,C**. Enrichment for common genetic associations with schizophrenia or ASD in groups of genes defined by mRNA localization and synaptic function. Displayed is the effect size (Beta) ± standard error from MAGMA competitive gene set association analysis. *P*-values denote significance of effect size comparisons between locally and distally translated synaptic transcripts in z-tests. “Localized mRNA: synaptic”: genes annotated to SynGO:*synapse* and enriched in the local transcriptome or translatome. “Non-localized mRNA: synaptic”: genes annotated to SynGO:*synapse* not enriched in the local transcriptome or translatome. “Localized mRNA: non-synaptic”: genes enriched in the local transcriptome or translatome but not annotated to SynGO:*synapse*.

#### FMRP targets

Pyramidal neuron FMRP-bound mRNA targets were taken from a study of RNA:protein cross-linking immunoprecipitation (CLIP) in mouse hippocampal CA1, in which FMRP was conditionally tagged using Cre-lox driven by a *Camk2a* promotor (32). Mouse samples were taken at 28-32 postnatal days. FMRP targets with a CLIP score > 1 were included (*stringent* and *high-binding* targets (32)). These 1265 FMRP-bound mRNAs were converted from mouse RefSeq mRNA IDs to human Ensembl gene IDs. 1242 FMRP targets remained after removal of genes with zero or multiple human homologs.

We acquired subcellular FMRP binding statistics, taken from pyramidal neuron dendrites and cell bodies separately, from published data by Hale and colleagues (30). Hippocampal slices from *Camk2a*-Cre-driven FMRP conditionally tagged adult (6-10 weeks) mice were microdissected into neuropil and cell body regions of CA1 before CLIP to purify FMRP-bound mRNA from these specific cellular compartments. Mouse RefSeq mRNA IDs for 5614 genes with CLIP scores were converted to 5303 human Ensembl IDs. CLIP scores were taken forward for gene property analysis. For gene set analysis, genes were ranked by their CLIP scores and the top 5300 were split into 25 bins of 212 genes.

### GWAS summary statistics

Common SNP associations with schizophrenia were determined from a recent genome-wide association study (GWAS) meta-analysis of 74,776 cases and 101,023 control individuals of European, East Asian, African American and Latino ancestry (2, 33) (primary meta-analysis). ASD GWAS summary statistics were taken from a meta-analysis of 18,381 individuals with ASD and 27,969 controls of European ancestry (4). SNPs with a minor allele frequency of less than 1% were excluded.

### Genetic association testing

Gene set and gene property association analyses were performed using multiple regression models in MAGMA v1.10 (34). GWAS SNPs were summarised to gene-wide *P*-values using the *SNP-wise Mean* model. A window of 35kb upstream and 10kb downstream was included to account for proximal regulatory regions. The Ensembl GRCh37 genome build was used for mapping and the European Phase 3 1000 Genomes Project reference data (35) was used to control for linkage disequilibrium. One-tailed competitive gene set analyses were performed to determine the strength of genetic associations with the phenotype in a set of genes compared to all remaining protein-coding genes, adjusting for potentially confounding effects of gene size, SNP density, and variations in sample size between SNPs. To compare the enrichment for associations between two non-overlapping or partially overlapping gene sets, a z-test of beta values was used. To compare the association between two sets of genes where one is a subset of the other, the smaller set was re-tested, and the larger set was added to the model as a conditional variable (34). MAGMA gene property analyses were used to test if continuous gene-level variables (e.g. FMRP CLIP scores) are related to stronger enrichment for genetic associations. Gene property analyses were two-tailed. Multiple independent tests were controlled for by adjusting *P*-values using the Bonferroni method.

## Results

### Genetic association with schizophrenia and ASD of synaptic genes split by mRNA localization

In competitive tests against all protein-coding genes, schizophrenia associations were enriched both in synaptic genes encoding mRNAs localized to cortical synaptoneurosomes (28) (β = 0.37, Bonferroni adjusted *P (P.adj)* = 5.4×10^-9^) and in synaptic genes encoding non-localized mRNAs (β = 0.055, *P.adj* = 0.030). However, from a comparison of effect sizes, we observed that localized synaptic mRNAs exhibited much stronger enrichment than non-localized synaptic mRNAs (Z = 3.2, *P* = 6.2×10^-4^) (Figure 1B). This relationship did not generalise to all transcripts localized to synaptoneurosomes, since localized transcripts without synaptic functions were depleted for associations with schizophrenia in comparison to those with synaptic functions (Z = -4.7, *P* = 1.4×10^-6^). Presynaptic and postsynaptic subsets of the local transcriptome exhibited no significant differences in association with schizophrenia (Supplementary Figure 2).

The enrichment for schizophrenia associations among local synaptic transcripts was reflected in repeated analyses using ribosome-bound mRNAs in dendrites from hippocampal pyramidal neurons (30) (Figure 1B). Genes encoding localized synaptic transcripts were more strongly associated than the remaining synaptic genes (Z = 2.2, *P* = 0.013) supporting the view that schizophrenia-related variants from GWAS preferentially impact the local synaptic translatome. Through repeated analyses using alternative synaptic gene annotations obtained from the GO database (27), we observed the same enrichment for schizophrenia associations in local synaptic transcripts (Supplementary Figure 3), indicating that this result is robust to variation in the definitions used for synaptic functioning.

In contrast, genetic associations with ASD were not significantly enriched in synaptic mRNAs localized to cortical synaptoneurosomes (28) (β = 0.10, *P.adj* = 0.061) or pyramidal neuron dendritic ribosomes (30) (β = 0.12, *P.adj* = 0.15), nor was it enriched in the remaining non-localized sets of synaptic genes which exhibited comparable effect sizes (Figure 1C).

The sets of localized mRNAs identified in studies by Ouwenga et al and Hale et al were captured in unstimulated tissues (28, 30). Since local protein synthesis is a key mechanism in activity-dependent synaptic processes (12, 36, 37) and the association of mRNAs with ribosomes is altered following stimulation (31), we performed an additional analysis to test whether transcripts bound to localized ribosomes following memory stimulation are enriched for genetic associations with schizophrenia and ASD. Synaptic genes encoding ribosome-bound mRNAs in hippocampal pyramidal neuron dendrites following a novel experience, consisting of a contextual fear conditioning trial (31), were enriched for associations with both schizophrenia and ASD (Schizophrenia: β = 0.36, *P.adj* = 3.0×10^-4^; ASD: β = 0.27, *P.adj* = 0.0011). These associations were stronger than the remaining synaptic genes (schizophrenia: Z = 1.7, *P* = 0.048; ASD: Z = 2.8, *P* = 0.0028) (Figure 1B, C). The same gene set was enriched for schizophrenia and ASD associations in comparison to the remaining local translatome (schizophrenia: Z = 2.4, *P* = 0.0081; ASD: Z = 2.7, *P* = 0.0039), showing that these relationships did not extend to all mRNAs binding to dendritic ribosomes after stimulation.

To determine if the selective enrichment of ASD associations in the activity-induced synaptic translatome is specific to the dendritic compartment, we performed further association analyses on mRNAs bound to ribosomes in the soma of the same neurons after contextual fear conditioning. Synaptic genes encoding transcripts in this somatic translatome were not enriched for ASD associations compared to all protein-coding genes (β = 0.10, *P.adj* = 0.062) or to the remaining synaptic genes (Z = 0.82, *P* = 0.21).

### Independence of genetic associations in the local synaptic translatome and FMRP targets

We hypothesised that the genetic associations with schizophrenia and ASD attributed to the local synaptic translatome are captured by the associations attributed to mRNA targets of FMRP. FMRP targets derived from hippocampal pyramidal neurons (32) were enriched for genetic associations with schizophrenia (β = 0.29, *P* = 2.0×10^-15^) and ASD (β = 0.091, *P* = 6.5×10^-4^). On average, 27.7% of FMRP targets overlapped with localized transcripts and 18.0% of FMRP targets overlapped with SynGO:*synapse* annotations (Supplementary Figure 1B). Conditioning on FMRP targets resulted in only minor reductions in the association of localized synaptic transcripts with schizophrenia or ASD (Figure 2A). Furthermore, localized subsets of FMRP targets were no more strongly enriched for schizophrenia or ASD associations than FMRP targets as a whole (Figure 2B). Taken together, these results suggest that the local synaptic translatome captures genetic association with these disorders that is independent of the association conferred through FMRP targets.

**Figure 2.**
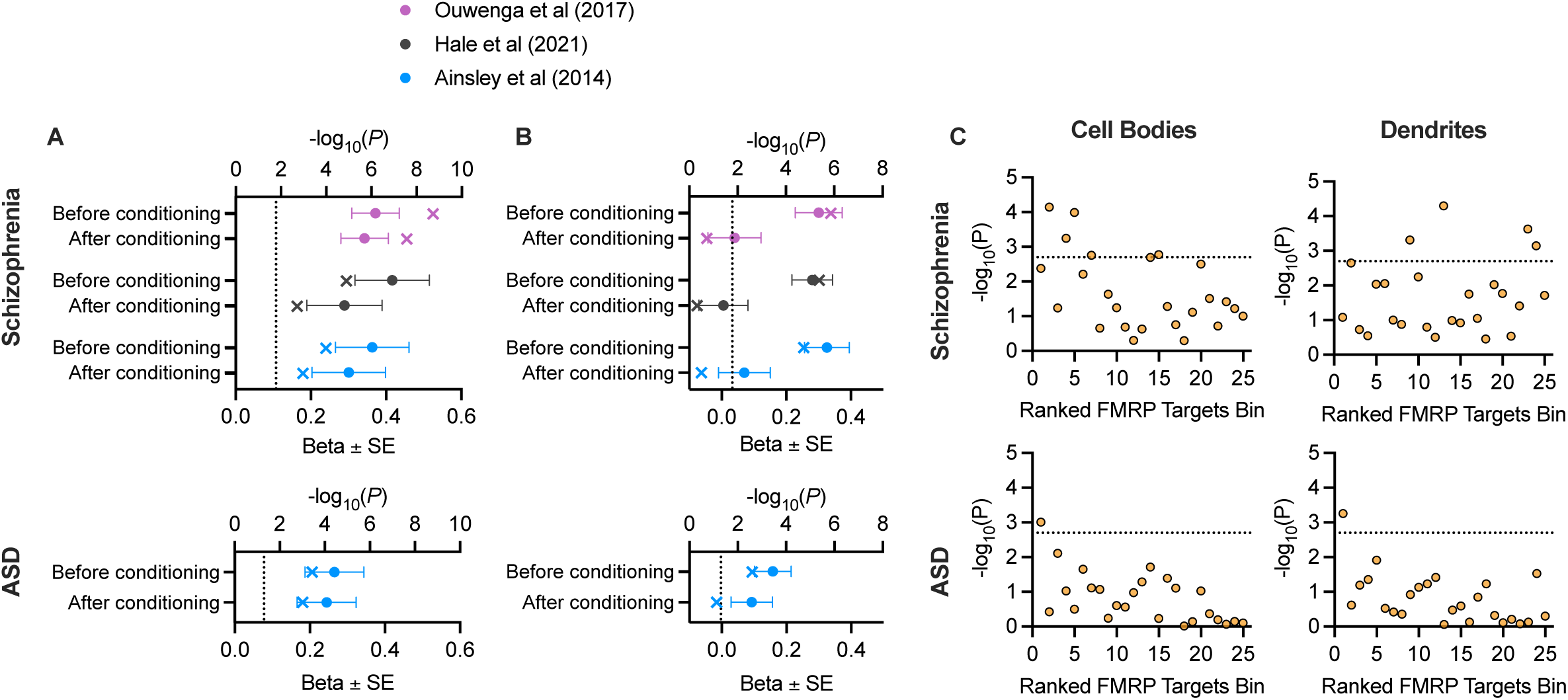
**A.** Enrichment of localized synaptic transcripts for common genetic associations with schizophrenia or ASD before and after conditioning on FMRP targets (32). Circles are the effect sizes (Beta) ± standard error in MAGMA competitive gene set association analysis. Crosses indicate -log_10_(*P*-value) for each test. The dotted lines indicate the threshold for significance (for schizophrenia, after correcting for 3 tests using the Bonferroni method). **B.** Enrichment of localized FMRP targets for common genetic associations with schizophrenia or ASD before and after conditioning on all FMRP targets. Displayed is the effect size (Beta) ± standard error. Crosses indicate -log_10_(*P*-value) for each test. **C**. Schizophrenia and ASD association of gene sets ranked by FMRP binding confidence in pyramidal neuron cell bodies and dendrites. Genes were ranked by CLIP score and divided into 25 bins of 212 genes. Each bin was subjected to competitive gene set association analysis in MAGMA. Displayed is -log_10_(*P*-value) for each test. The dotted line indicates the threshold for significance after adjusting for 25 tests using the Bonferroni method.

To investigate this further, we explored the relationship between schizophrenia and ASD genetic association and FMRP-mRNA binding CLIP scores obtained in hippocampal pyramidal neuron dendrites and cell bodies separately (30). Schizophrenia association was related to FMRP CLIP scores derived from somata (β = 0.045, *P* = 5.7×10^-4^) but not with those from dendrites (β = -0.023, *P* = 0.87). On the other hand, ASD association was significantly related to FMRP CLIP scores from both soma (β = 0.023, *P* = 0.012) and dendrites (β = 0.036, *P* = 0.0087). Accordingly, bins of genes with higher FMRP CLIP scores in cell bodies were enriched for associations with both schizophrenia and ASD, whilst bins with high dendritic FMRP CLIP scores were only enriched for ASD associations (Figure 2C). Hence, mRNA-FMRP binding in the somata, but not in dendrites, was related to schizophrenia genetic risk. This provides additional evidence that schizophrenia genetic risk conferred through FMRP targets is separate from that conferred through the local synaptic translatome.

## Discussion

Genes annotated to synaptic functions are strongly implicated in risk conferred to schizophrenia and ASD (2, 3, 7, 8, 10). We tested whether the common variant associations attributed to the synapse in these disorders are over-represented within genes encoding mRNAs localized to dendrites, available for rapid synthesis in response to synaptic activity. Schizophrenia associations were enriched in localized synaptic transcripts identified from cortical synaptoneurosomes or hippocampal dendritic ribosomes, including those captured following memory stimulation. ASD associations were enriched in localized synaptic transcripts only following memory stimulation. In each case, the genetic associations captured by localized synaptic transcripts did not explain the enrichment of associations in FMRP targets.

Our results support the hypothesis that synaptic pathways responsible for time- and spatially-sensitive molecular processes are particularly impacted by risk variants associated with schizophrenia. These processes may include those required for synaptic plasticity in adulthood, such as long-term potentiation, and those responsible for establishing early synaptic connectivity and maturation during development. Both mature and developmental plasticity pathways have been previously implicated in risk for schizophrenia (7). The present findings now highlight a subset of these pathways which may be particularly adapted to rapid, compartment-specific activity-dependent functions.

The pattern of association with ASD was somewhat different to that for schizophrenia, with enrichment for associations only being observed in genes encoding the local synaptic translatome of hippocampal pyramidal neurons from mice exposed to a contextual fear conditioning trial (31). Localized synaptic transcripts obtained from the same cellular compartment in mice without stimulation (30), or from cortical synaptoneurosomes (28), were not enriched for ASD associations. Our results suggest that ASD risk is enriched in a subset of synaptic genes encoding mRNAs which rapidly bind to dendritic ribosomes during neuronal stimulation in the hippocampus, such as that accompanying memory acquisition. Since this relationship between ASD association and stimulation-induced ribosome binding was not reflected in the cell body, we conclude that specifically dendrite-localized mRNA translation after stimulation was responsible for the enrichment of genetic associations among synaptic genes. More broadly, our results are consistent with previous reports linking activity-dependent pathways to ASD (38, 39). It is important to note that the ASD GWAS is substantially less powered than the schizophrenia GWAS which may influence comparisons between the disorders. Furthermore, comparisons of transcriptomic datasets adopted by our study may be affected by undefined variables from methodological differences between the original studies.

Our investigation focused on the local translatome in excitatory glutamatergic neurons and may not reflect patterns of genetic risk in other neuronal subtypes (or non-neuronal cells). Previous evidence shows that inhibitory neurons, including cortical interneurons and medium spiny neurons, and their synaptic complexes may play an important role in conferring risk to schizophrenia (2, 40, 41). However, recent work comparing local transcriptomes between multiple cell types reported few differences in transcripts observed in dendritic fractions of glutamatergic and GABAergic neurons (42) or in synaptosomes from glutamatergic and dopaminergic neurons (43). Therefore, impacts of psychiatric risk variants on localized synaptic transcripts may be common to multiple types of neurons.

Local translation plays an important role in both developing and mature neurons (12), but our study focused only on transcriptomic data from mice of at least 21 postnatal days. Localization of mRNAs in neurites differs by developmental stage (44, 45) in response to altered spatial and temporal dependencies, and this divergence is likely amplified by stimulation. Local translatomes in developing neurons may capture additional genetic risk for neuropsychiatric and neurodevelopmental disorders, conferring effects as the brain matures. Establishing precisely at which developmental stages, and under what conditions, localized transcripts confer genetic risk to these disorders could help refine targets for treatment.

Despite the proposed role of FMRP as a key regulator of local translation of synaptic mRNAs (46) and the association of its targets with schizophrenia and ASD (15, 19, 20), we observed that the genetic risk attributed to the local synaptic translatome was independent of FMRP targets in both disorders. Furthermore, the localization of FMRP targets to dendrites was not related to increased genetic association with either disorder. In the case of schizophrenia, only FMRP binding in the cell body was related to genetic risk. Soma-derived FMRP targets are enriched for synaptic functional annotations (30), but those contributing to psychiatric risk, particularly for schizophrenia, may encode proteins translated prior to transport, or bound for reasons unrelated to translational regulation. In two previous studies of altered translation or ribosome occupancy following loss of FMRP in retinal ganglionic cells (47) or Cath.a-differentiated (CAD) neurons (48), affected transcripts were not enriched for synaptic functions. FMRP is reported to have functions beyond translational repression, including RNA transport (48), regulation of RNA stability (49), and RNA splicing (50, 51), for which it may target distinct pools of transcripts (48). It has been suggested that FMRP preferentially targets and stabilizes long transcripts encoding complex proteins required for synaptic development and plasticity (52–54), thereby tagging a set of genes characterized by an intersection of function and regulatory requirements. The FMRP binding data used in the present study were obtained from mature hippocampal tissue at rest. Under stimulation, or at alternative developmental stages, the transcripts targeted, or where in the cell they are bound, may differ, and therefore the relationships between FMRP binding and genetic associations with schizophrenia and ASD may also differ.

Aside from FMRP, additional, functionally related RBPs have genetic links to schizophrenia and ASD. Transcripts bound by RBPs of the cytoplasmic polyadenylation element binding (CPEB) family are also enriched for common genetic associations with schizophrenia and ASD (55, 56). Furthermore, rare variants in RBP genes, *CSDE1* and *RBFOX1*, have been linked to ASD (57–59), whilst common variation in *RBFOX1* is associated with schizophrenia and other psychiatric conditions (2, 59). Critically, the binding targets of CPEBs, CSDE1 and RBFOX1 are enriched for FMRP targets and share similar functional representation, including the regulation of neuronal development and synaptic plasticity (55, 57, 60, 61).

To conclude, we provide evidence that the degree of synaptic gene enrichments for common genetic associations with schizophrenia and ASD depends on the subcellular localizations in neurons of the cognate encoded mRNAs, and suggest that those that locally translated in dendritic compartments are particularly relevant to the pathogenesis of these disorders. However, despite FMRP playing a role in local translation, and the fact that genes encoding mRNA targets of FMRP are also enriched for genetic associations to schizophrenia and ASD, those associations are independent of localized synaptic mRNAs. In schizophrenia, the subset of mRNAs bound by FMRP in the cell body rather than proximal to synapses are associated with genetic risk. These results imply that the pathophysiological effects on schizophrenia and ASD indexed by FMRP binding function are unlikely to be related to local translation of those transcripts. Further work examining RNA regulation across neurodevelopment and states of activation could help to elucidate precisely which mechanisms are key to the genetic risk conferred to a range of different neurodevelopmental and neuropsychiatric disorders.

## Data Availability

All data produced in the present study are available upon reasonable request to the authors

## Acknowledgements

This work was funded by a UKRI Career Development Award to NEC (MR/W017156/1). The study was supported by the National Institute for Health and Care Research Exeter Biomedical Research Centre. The views expressed are those of the author(s) and not necessarily those of the NIHR or the Department of Health and Social Care. We thank the Psychiatric Genomics Consortium for making available genomics datasets used in this study.

## Supplementary Information

**Supplementary Figure 1.**
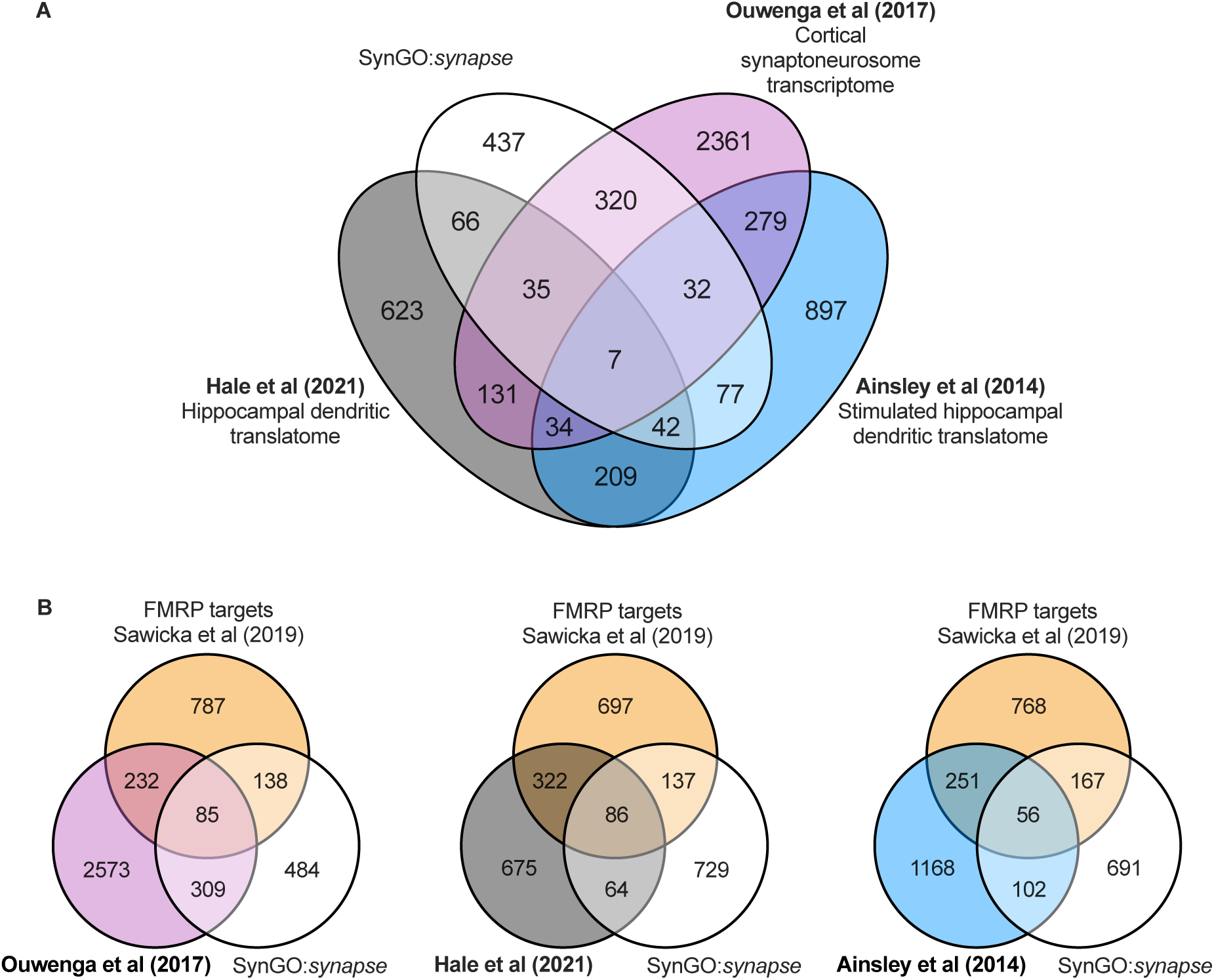
**A.** Intersection of localized mRNA transcripts with SynGO:*synapse* gene annotations. **B.** Intersection of localized mRNA transcripts with FMRP targets and SynGO:*synapse* annotations. Linked to Figures 2A and 2B.

**Supplementary Figure 2.**
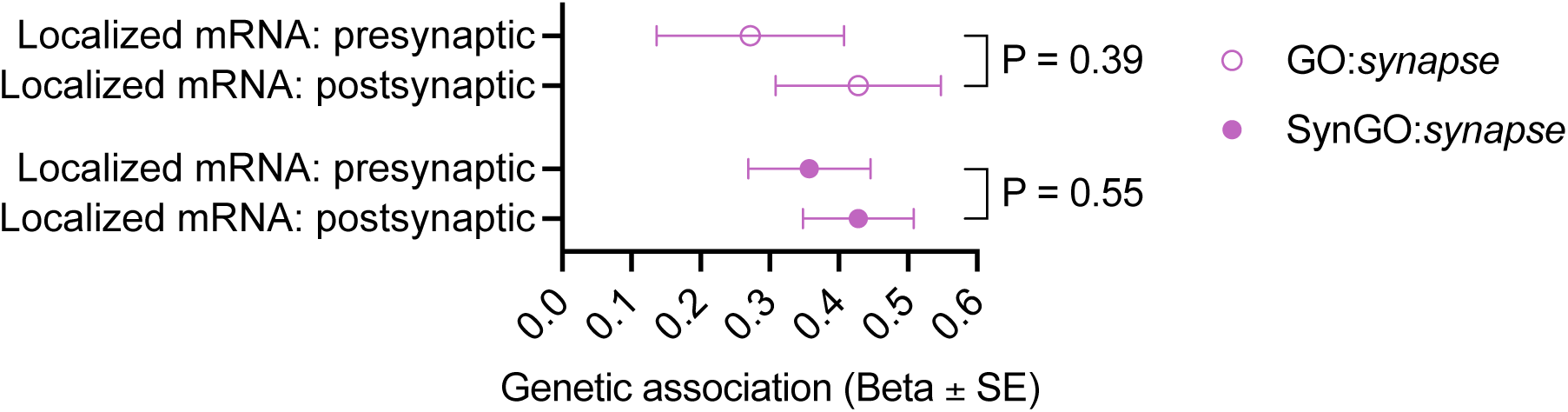
Comparison of genetic association with schizophrenia of the local synaptic transcriptome (Ouwenga et al, 2017) from presynaptic or postsynaptic compartments. Analyses were repeated using functional annotations from SynGO:*synapse* or GO:*synapse*. Displayed is the effect size (Beta) ± standard error from MAGMA competitive gene set association analysis of common genetic variation associated with schizophrenia through GWAS. *P*-values represent significance in z-test of beta values obtained from analyses of presynaptic and postsynaptic subsets.

**Supplementary Figure 3.**
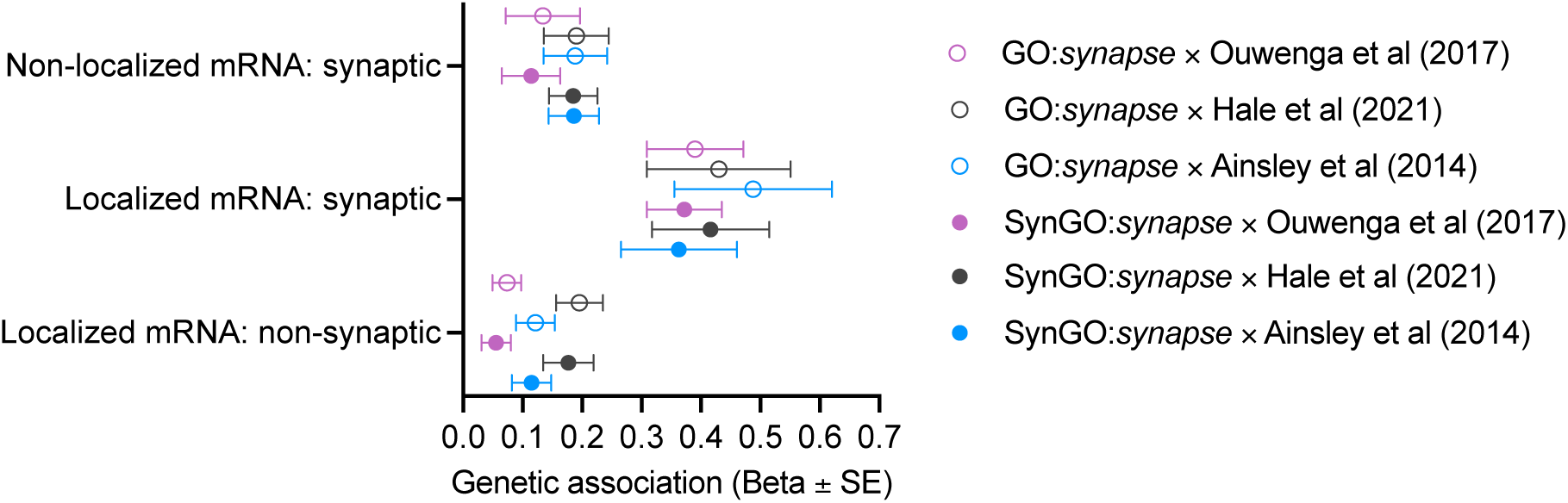
Enriched genetic associations with schizophrenia in localized synaptic transcripts is robust to variation in synaptic functional annotations. Localized transcripts identified in each of three studies were intersected with either GO:*synapse* annotations or SynGO:*synapse* annotations. “Localized mRNA: synaptic”: genes annotated to SynGO:*synapse* and enriched in the local transcriptome or translatome. “Non-localized mRNA: synaptic”: genes annotated to SynGO:*synapse* not enriched in the local transcriptome or translatome. “Localized mRNA: non-synaptic”: genes enriched in the local transcriptome or translatome but not annotated to SynGO:*synapse*. Displayed is the effect size (Beta) ± standard error from MAGMA competitive gene set association analysis of common variation associated with schizophrenia.

## References

1. GBD 2019 Mental Disorders Collaborators (2022): Global, regional, and national burden of 12 mental disorders in 204 countries and territories, 1990-2019: a systematic analysis for the global burden of disease study 2019. Lancet Psychiatry. 9(2): 137–50.

2. Trubetskoy V, Pardiñas AF, Qi T, Panagiotaropoulou G, Awasthi S, Bigdeli TB, et al. (2022): Mapping genomic loci implicates genes and synaptic biology in schizophrenia. Nature. 604(7906): 502–8.

3. Singh T, Poterba T, Curtis D, Akil H, Al Eissa M, Barchas JD, et al. (2022): Rare coding variants in ten genes confer substantial risk for schizophrenia. Nature. 604(7906): 509–16.

4. Grove J, Ripke S, Als TD, Mattheisen M, Walters RK, Won H, et al. (2019): Identification of common genetic risk variants for autism spectrum disorder. Nat. Genet. 51(3): 431–44.

5. Wang T, Kim CN, Bakken TE, Gillentine MA, Henning B, Mao Y, et al. (2022): Integrated gene analyses of de novo variants from 46,612 trios with autism and developmental disorders. Proc. Natl. Acad. Sci. USA. 119(46): e2203491119.

6. Satterstrom FK, Kosmicki JA, Wang J, Breen MS, De Rubeis S, An J-Y, et al. (2020): Large-scale exome sequencing study implicates both developmental and functional changes in the neurobiology of autism. Cell. 180(3): 568–584.e23.

7. Hall J, Bray NJ (2022): Schizophrenia genomics: convergence on synaptic development, adult synaptic plasticity, or both? Biol. Psychiatry. 91(8): 709–17.

8. Hansel C (2019): Deregulation of synaptic plasticity in autism. Neurosci. Lett. 688: 58–61.

9. Klein ME, Monday H, Jordan BA (2016): Proteostasis and rna binding proteins in synaptic plasticity and in the pathogenesis of neuropsychiatric disorders. Neural Plast. 2016: 3857934.

10. Bourgeron T (2015): From the genetic architecture to synaptic plasticity in autism spectrum disorder. Nat. Rev. Neurosci. 16(9): 551–63.

11. Forsyth JK, Lewis DA (2017): Mapping the consequences of impaired synaptic plasticity in schizophrenia through development: an integrative model for diverse clinical features. Trends Cogn. Sci. (Regul. Ed.). 21(10): 760–78.

12. Holt CE, Martin KC, Schuman EM (2019): Local translation in neurons: visualization and function. Nat. Struct. Mol. Biol. 26(7): 557–66.

13. Stefani G, Fraser CE, Darnell JC, Darnell RB (2004): Fragile x mental retardation protein is associated with translating polyribosomes in neuronal cells. J. Neurosci. 24(33): 7272–76.

14. Darnell JC, Van Driesche SJ, Zhang C, Hung KYS, Mele A, Fraser CE, et al. (2011): FMRP stalls ribosomal translocation on mrnas linked to synaptic function and autism. Cell. 146(2): 247–61.

15. Clifton NE, Rees E, Holmans PA, Pardiñas AF, Harwood JC, Di Florio A, et al. (2021): Genetic association of fmrp targets with psychiatric disorders. Mol. Psychiatry. 26(7): 2977–90.

16. Pardiñas AF, Holmans P, Pocklington AJ, Escott-Price V, Ripke S, Carrera N, et al. (2018): Common schizophrenia alleles are enriched in mutation-intolerant genes and in regions under strong background selection. Nat. Genet. 50(3): 381–89.

17. Iossifov I, O’Roak BJ, Sanders SJ, Ronemus M, Krumm N, Levy D, et al. (2014): The contribution of de novo coding mutations to autism spectrum disorder. Nature. 515(7526): 216– 21.

18. De Rubeis S, He X, Goldberg AP, Poultney CS, Samocha K, Cicek AE, et al. (2014): Synaptic, transcriptional and chromatin genes disrupted in autism. Nature. 515(7526): 209–15.

19. Jansen A, Dieleman GC, Smit AB, Verhage M, Verhulst FC, Polderman TJC, et al. (2017): Gene-set analysis shows association between fmrp targets and autism spectrum disorder. Eur. J. Hum. Genet. 25(7): 863–68.

20. Zhou X, Feliciano P, Shu C, Wang T, Astrovskaya I, Hall JB, et al. (2022): Integrating de novo and inherited variants in 42,607 autism cases identifies mutations in new moderate-risk genes. Nat. Genet. 54(9): 1305–19.

21. Butler MG (2017): Clinical and genetic aspects of the 15q11.2 bp1-bp2 microdeletion disorder. J. Intellect. Disabil. Res. 61(6): 568–79.

22. Clifton NE, Thomas KL, Wilkinson LS, Hall J, Trent S (2020): FMRP and cyfip1 at the synapse and their role in psychiatric vulnerability. Complex Psychiatry. 6(1–2): 5–19.

23. Rees E, Creeth HDJ, Hwu H-G, Chen WJ, Tsuang M, Glatt SJ, et al. (2021): Schizophrenia, autism spectrum disorders and developmental disorders share specific disruptive coding mutations. Nat. Commun. 12(1): 5353.

24. Autism Spectrum Disorders Working Group of The Psychiatric Genomics Consortium (2017): Meta-analysis of gwas of over 16,000 individuals with autism spectrum disorder highlights a novel locus at 10q24.32 and a significant overlap with schizophrenia. Mol. Autism. 8: 21.

25. McCarthy SE, Gillis J, Kramer M, Lihm J, Yoon S, Berstein Y, et al. (2014): De novo mutations in schizophrenia implicate chromatin remodeling and support a genetic overlap with autism and intellectual disability. Mol. Psychiatry. 19(6): 652–58.

26. Koopmans F, van Nierop P, Andres-Alonso M, Byrnes A, Cijsouw T, Coba MP, et al. (2019): SynGO: an evidence-based, expert-curated knowledge base for the synapse. Neuron. 103(2): 217–234.e4.

27. The Gene Ontology Consortium (2017): Expansion of the gene ontology knowledgebase and resources. Nucleic Acids Res. 45(D1): D331–38.

28. Ouwenga R, Lake AM, O’Brien D, Mogha A, Dani A, Dougherty JD (2017): Transcriptomic analysis of ribosome-bound mrna in cortical neurites in vivo. J. Neurosci. 37(36): 8688–8705.

29. Smedley D, Haider S, Durinck S, Pandini L, Provero P, Allen J, et al. (2015): The biomart community portal: an innovative alternative to large, centralized data repositories. Nucleic Acids Res. 43(W1): W589–98.

30. Hale CR, Sawicka K, Mora K, Fak JJ, Kang JJ, Cutrim P, et al. (2021): FMRP regulates mrnas encoding distinct functions in the cell body and dendrites of ca1 pyramidal neurons. Elife. 10:

31. Ainsley JA, Drane L, Jacobs J, Kittelberger KA, Reijmers LG (2014): Functionally diverse dendritic mrnas rapidly associate with ribosomes following a novel experience. Nat. Commun. 5: 4510.

32. Sawicka K, Hale CR, Park CY, Fak JJ, Gresack JE, Van Driesche SJ, et al. (2019): FMRP has a cell-type-specific role in ca1 pyramidal neurons to regulate autism-related transcripts and circadian memory. Elife. 8:

33. Bigdeli TB, Genovese G, Georgakopoulos P, Meyers JL, Peterson RE, Iyegbe CO, et al. (2020): Contributions of common genetic variants to risk of schizophrenia among individuals of african and latino ancestry. Mol. Psychiatry. 25(10): 2455–67.

34. de Leeuw CA, Mooij JM, Heskes T, Posthuma D (2015): MAGMA: generalized gene-set analysis of gwas data. PLoS Comput. Biol. 11(4): e1004219.

35. 1000 Genomes Project Consortium, Auton A, Brooks LD, Durbin RM, Garrison EP, Kang HM, et al. (2015): A global reference for human genetic variation. Nature. 526(7571): 68–74.

36. Monday HR, Kharod SC, Yoon YJ, Singer RH, Castillo PE (2022): Presynaptic fmrp and local protein synthesis support structural and functional plasticity of glutamatergic axon terminals. Neuron. 110(16): 2588–2606.e6.

37. Bradshaw KD, Emptage NJ, Bliss TVP (2003): A role for dendritic protein synthesis in hippocampal late ltp. Eur. J. Neurosci. 18(11): 3150–52.

38. Boulting GL, Durresi E, Ataman B, Sherman MA, Mei K, Harmin DA, et al. (2021): Activity-dependent regulome of human gabaergic neurons reveals new patterns of gene regulation and neurological disease heritability. Nat. Neurosci. 24(3): 437–48.

39. Ebert DH, Greenberg ME (2013): Activity-dependent neuronal signalling and autism spectrum disorder. Nature. 493(7432): 327–37.

40. Pocklington AJ, Rees E, Walters JTR, Han J, Kavanagh DH, Chambert KD, et al. (2015): Novel findings from cnvs implicate inhibitory and excitatory signaling complexes in schizophrenia. Neuron. 86(5): 1203–14.

41. Skene NG, Bryois J, Bakken TE, Breen G, Crowley JJ, Gaspar HA, et al. (2018): Genetic identification of brain cell types underlying schizophrenia. Nat. Genet. 50(6): 825–33.

42. Perez JD, Dieck ST, Alvarez-Castelao B, Tushev G, Chan IC, Schuman EM (2021): Subcellular sequencing of single neurons reveals the dendritic transcriptome of gabaergic interneurons. Elife. 10:

43. Hobson BD, Kong L, Angelo MF, Lieberman OJ, Mosharov EV, Herzog E, et al. (2022): Subcellular and regional localization of mrna translation in midbrain dopamine neurons. Cell Rep. 38(2): 110208.

44. Shigeoka T, Jung H, Jung J, Turner-Bridger B, Ohk J, Lin JQ, et al. (2016): Dynamic axonal translation in developing and mature visual circuits. Cell. 166(1): 181–92.

45. Poulopoulos A, Murphy AJ, Ozkan A, Davis P, Hatch J, Kirchner R, et al. (2019): Subcellular transcriptomes and proteomes of developing axon projections in the cerebral cortex. Nature. 565(7739): 356–60.

46. Darnell JC, Klann E (2013): The translation of translational control by fmrp: therapeutic targets for fxs. Nat. Neurosci. 16(11): 1530–36.

47. Jung J, Ohk J, Kim H, Holt CE, Park HJ, Jung H (2022): MRNA transport, translation, and decay in adult mammalian central nervous system axons. Neuron

48. Goering R, Hudish LI, Guzman BB, Raj N, Bassell GJ, Russ HA, et al. (2020): FMRP promotes rna localization to neuronal projections through interactions between its rgg domain and g-quadruplex rna sequences. Elife. 9:

49. Shu H, Donnard E, Liu B, Jung S, Wang R, Richter JD (2020): FMRP links optimal codons to mrna stability in neurons. Proc. Natl. Acad. Sci. USA. 117(48): 30400–411.

50. Shah S, Molinaro G, Liu B, Wang R, Huber KM, Richter JD (2020): FMRP control of ribosome translocation promotes chromatin modifications and alternative splicing of neuronal genes linked to autism. Cell Rep. 30(13): 4459–4472.e6.

51. Zhou L-T, Ye S-H, Yang H-X, Zhou Y-T, Zhao Q-H, Sun W-W, et al. (2017): A novel role of fragile x mental retardation protein in pre-mrna alternative splicing through rna-binding protein 14. Neuroscience. 349: 64–75.

52. Seo SS, Louros SR, Anstey N, Gonzalez-Lozano MA, Harper CB, Verity NC, et al. (2022): Excess ribosomal protein production unbalances translation in a model of fragile x syndrome. Nat. Commun. 13(1): 3236.

53. Flanagan K, Baradaran-Heravi A, Yin Q, Dao Duc K, Spradling AC, Greenblatt EJ (2022): FMRP-dependent production of large dosage-sensitive proteins is highly conserved. Genetics. 221(4):

54. Greenblatt EJ, Spradling AC (2018): Fragile x mental retardation 1 gene enhances the translation of large autism-related proteins. Science. 361(6403): 709–12.

55. Ollà I, Pardiñas AF, Parras A, Hernández IH, Santos-Galindo M, Picó S, et al. (2023): Pathogenic mis-splicing of cpeb4 in schizophrenia. Biol. Psychiatry

56. Parras A, Anta H, Santos-Galindo M, Swarup V, Elorza A, Nieto-González JL, et al. (2018): Autism-like phenotype and risk gene mrna deadenylation by cpeb4 mis-splicing. Nature. 560(7719): 441–46.

57. Guo H, Li Y, Shen L, Wang T, Jia X, Liu L, et al. (2019): Disruptive variants of csde1 associate with autism and interfere with neuronal development and synaptic transmission. Sci. Adv. 5(9): eaax2166.

58. Bacchelli E, Cameli C, Viggiano M, Igliozzi R, Mancini A, Tancredi R, et al. (2020): An integrated analysis of rare cnv and exome variation in autism spectrum disorder using the infinium psycharray. Sci. Rep. 10(1): 3198.

59. O’Leary A, Fernàndez-Castillo N, Gan G, Yang Y, Yotova AY, Kranz TM, et al. (2022): Behavioural and functional evidence revealing the role of rbfox1 variation in multiple psychiatric disorders and traits. Mol. Psychiatry

60. Ivshina M, Lasko P, Richter JD (2014): Cytoplasmic polyadenylation element binding proteins in development, health, and disease. Annu. Rev. Cell Dev. Biol. 30: 393–415.

61. Lee J-A, Damianov A, Lin C-H, Fontes M, Parikshak NN, Anderson ES, et al. (2016): Cytoplasmic rbfox1 regulates the expression of synaptic and autism-related genes. Neuron. 89(1): 113–28.

